# Label-Free Threshold Selection for Out-of-Distribution Detection in Liver CT Segmentation

**DOI:** 10.64898/2026.08.20.26360809

**Authors:** Marshall Nielsen, Austin Castelo, Mais Altaie, Jeddy Bennett, Ajith Anthony, Noreen S. Siddiqi, Aashish Chandra Gupta, Kristy K. Brock, McKell Woodland

**Affiliations:** Department of Imaging Physics, The University of Texas MD Anderson Cancer Center, Houston TX 77030, USA; Department of Interventional Radiology, The University of Texas MD Anderson Cancer Center, Houston TX 77030, USA; Department of Statistics, Brigham Young University, Provo UT 84602, USA; Department of Mathematics, Brigham Young University, Provo UT 84602, USA

**Keywords:** Out-of-distribution detection, Failure detection, Liver segmentation, Computed tomography, Threshold Selection, Pairwise Surface DSC

## Abstract

Reliable clinical deployment of automated liver segmentation requires mechanisms for detecting failures in rare and previously unseen scenarios. Achieving this goal requires an appropriately calibrated threshold that converts an out-of-distribution (OOD) score into a failure prediction. However, threshold calibration typically relies on expert-labeled failures, creating a substantial annotation burden when failures are rare. Building upon our prior work, which uses Pairwise Surface DSC scores as indicators of segmentation quality, we propose a label-free framework for calibrating OOD score thresholds. First, we fitted a log-*t* distribution to Pairwise Surface DSC scores from a validation set of 400 internal scans to approximate an in-distribution score distribution. New segmentations were assigned significance scores based on their extremity under this fitted distribution and categorized into Low, Medium, and High Risk review groups using statistically principled cutoffs of 0.25 and 0.05. The fitted log-*t* distribution provided a strong fit to the observed scores and remained robust to moderate contamination by OOD cases. On an independent test set of 500 internal and external scans, the combined Medium and High Risk categories achieved 100% sensitivity and 79% specificity, whereas the High Risk category alone achieved 78% sensitivity and 96% specificity. These results indicate that clinically meaningful failure detection can be derived from unlabeled data. Our code is available at https://github.com/marshalln7/Label_Free_OOD_Threshold_Selection.

## 1 Introduction

Liver cancer is a leading cause of cancer-related mortality worldwide [1]. Radio-therapy is a common treatment modality that requires accurate delineation of tumors and surrounding organs at risk [2]. Traditionally performed manually, this segmentation process is increasingly being automated using deep learning methods to improve efficiency [3], consistency [4], and standardization.

Although AI-based segmentation methods perform well in most cases, they tend to fail when presented with image attributes that differ from those encountered during training [5]. To mitigate the risk of automation bias, out-of-distribution (OOD) detection has emerged as a mechanism for identifying cases that are likely to cause model failure [6]. Despite being motivated by the need to identify model failures in safety-critical settings, OOD detection has largely been evaluated using theoretical benchmarks, with relatively few studies assessing their effectiveness for identifying failure cases in real clinical environments [7].

Recently, Bennett et al. evaluated multiple OOD detection methods for a deployed liver CT segmentation model across diverse clinical datasets [8]. To determine an appropriate cutoff for the OOD score above which a segmentation would be predicted to be a failure, the authors optimized Youden’s J statistic on the validation set [9], which balances sensitivity and specificity. This threshold selection process was resource-intensive, requiring expert review of approximately 400 segmentations due to the relatively low prevalence of failure cases (13%).

As segmentation performance continues to improve and failures become increasingly rare, the annotation burden associated with threshold calibration is likely to grow, limiting the practical deployment of existing approaches, particularly in resource-constrained clinical settings. We investigate whether clinically meaningful OOD detection thresholds can be derived from validation data without labels and make the following contributions:

1. **Label-free threshold selection for OOD detection.** We introduce a method for selecting OOD detection thresholds by estimating the in-distribution of scores, eliminating the need for expert-annotated validation data, thereby reducing the cost of clinical deployment.
2. **Flexible reference distribution modeling.** We established that our Pair-wise Surface DSC scores were well described by a log-*t* distribution and further proposed an Ordered Quantile Normalization (ORQ)-based approach for label-free calibration when parametric assumptions are not satisfied.
3. **Empirical validation on real-world liver CT data.** Using the same clinically deployed segmentation model and multi-center liver CT datasets as Bennett et al. [8], the proposed label-free framework successfully identified all segmentation failures at the medium risk threshold, and successfully partitioned off cases with a high probability of being failures using the high risk threshold (45% positive predictive value).

## 2 Methods

Section 2.1 describes our validation and test datasets of liver CT scans. Section 2.2 introduces the clinically deployed segmentation model. Section 2.3 delineates the utilized OOD scoring techniques. Section 2.4 introduces our novel approach for selecting score thresholds in the absence of labels, which Figure 1 gives a high level overview of.

**Fig. 1.**
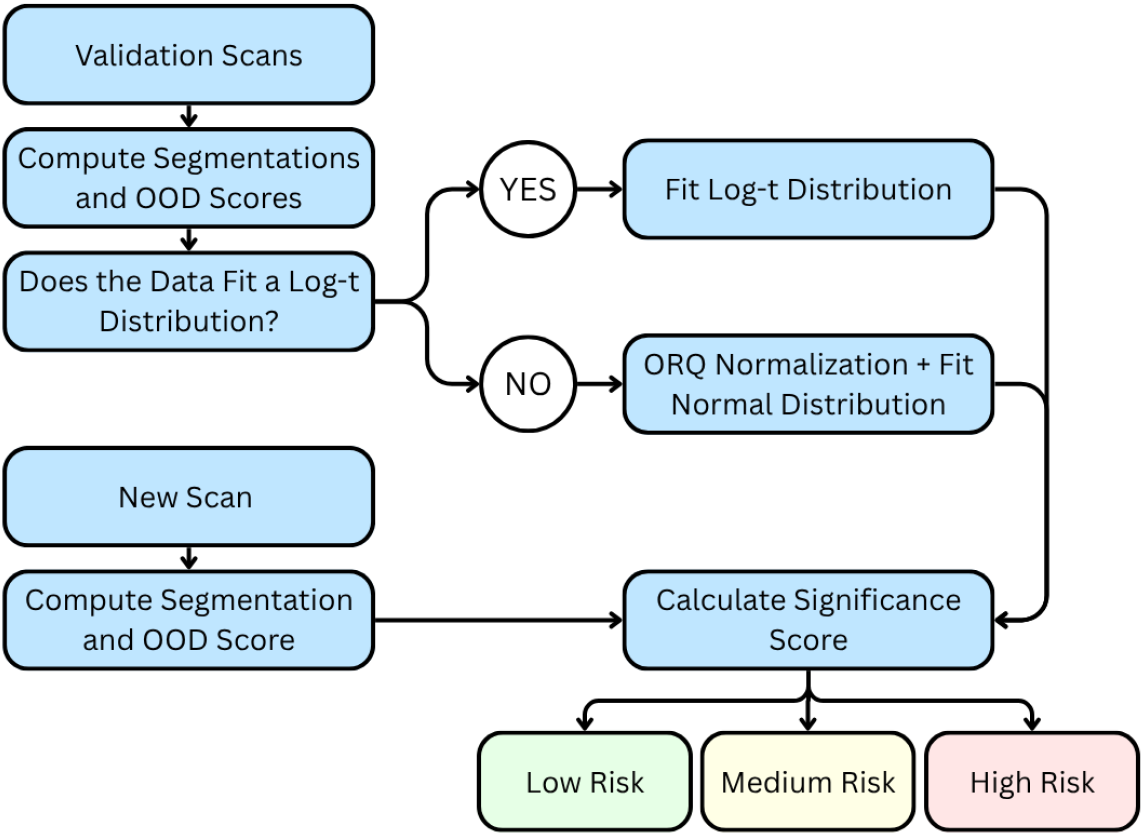
A flowchart depicting our proposed workflow.

### 2.1 Data

In this study, we used the validation and test cohorts from Bennett et al. [8], which were obtained from routine clinical workflows at The University of Texas MD Anderson Cancer Center under an approved Institutional Review Board protocol (PA18-0832), with informed consent waived due to the retrospective nature of the study. The validation cohort consisted of 400 liver CT scans (400 patients), with each segmentation labeled as a success or failure by a radiologist with 7 years of experience based on whether major edits (edits to *≥*10% of liver slices) would be required before clinical use. Although these labels were available for validation, they were not used to assign risk levels to test scans in this study; risk levels were determined solely from the distribution of OOD scores in the validation cohort.

Our test cohort consisted of 400 internal and 100 external liver CT scans from 500 unique patients. Three readers independently assigned five-point Likert ratings to each segmentation (1: unusable, 2: major edits with no time savings, 3: major edits with time savings, 4: minor edits, 5: no edits), with ratings of less than 4 considered failures, consistent with the validation set. The majority vote of the three success/failure assessments became the final label.

Our datasets contain two types of distribution shifts. First, there is a prior probability shift between the validation dataset (failure rate of 13.0%) and the test set (failure rate of 3.6%). This is attributable to the fact that the labels in the test set were a majority vote between three readers while the labels on the validation dataset came from a single reader. Second, an acquisition shift, as the 100 external test scans were sourced from over 70 sites across seven countries, whereas the validation dataset was sourced internally.

### 2.2 Segmentation Model

The liver CT segmentation model used in this study was also identical to that in Bennett et al. [8]. The model was developed using the 3D full-resolution nnU-Net framework [10] and trained on 2,840 CT scans using five-fold cross-validation [11]. At evaluation, the model achieved a mean Dice Similarity Coefficient (DSC) of 0.97 across internal and external datasets. The model had been clinically deployed prior to the start of this study.

### 2.3 OOD Scoring

The five segmentations produced by the five cross-validation models were compared in a pairwise fashion using Surface DSC [12] and DSC [13,14]. Surface DSC quantifies the proportion of surface agreement within a 2 mm tolerance, while DSC quantifies volumetric overlap, with higher agreement producing scores approaching one. The resulting ten values from each metric were averaged and subtracted from one to produce a single OOD score per segmentation, referred to as the Pairwise Surface DSC score [8] and Pairwise DSC score [15,7], respectively. We focused exclusively on pairwise techniques in this study because they were the two best-performing methods in Bennett et al. [8], with Pairwise Surface DSC demonstrating the best overall performance.

### 2.4 Threshold Selection

Once the validation scans were collected, segmented, and scored, we employed our new threshold selection process. To do this, the distribution of those scores was examined to see whether the observed scores were well described by a log-normal, beta, or log-*t* distribution using Q-Q plots. These candidate distributions were chosen because they can model a range of distributional shapes commonly observed in OOD scores, including skewness and heavy-tailed behavior. If none of the candidate distributions exhibited sufficiently linear Q-Q behavior, we employed a rank-based alternative. Specifically, ORQ normalization can be used to transform the observed scores into a normal distribution while preserving their rank ordering. We then fit the chosen distribution to the validation OOD scores. Once an appropriate reference distribution has been established, it can be used to evaluate new segmentations. For a given segmentation, its OOD score is compared against the approximated in-distribution to estimate a significance score, defined as the probability mass in the upper tail beyond the observed value. This significance score is then mapped to a risk category using predefined thresholds.

Segmentations are classified as Low Risk when their significance score exceeded 0.25, Medium Risk when the score fell between 0.05 and 0.25, and High Risk when the score was below 0.05. These thresholds were chosen apriori. As the Medium Risk category was purposed to capture all failures, we set its cutoff to the lower boundary of the highest quartile of the approximated in-distribution. The High Risk threshold was chosen to be 0.05, a widely accepted significance threshold in statistical hypothesis testing, to have a category with less false positives.

For the sake of comparison, the Youden’s index optimized cutoff score was also calculated for both Surface DSC and DSC. This was done by iterating over the data points in order and finding the inclusive threshold that yielded the highest Youden’s J statistic (J = Sensitivity + Specificity - 1), a process which required using the labels in the validation set. Our code is available at https://github.com/marshalln7/Label_Free_OOD_Threshold_Selection.

## 3 Experiments and Results

### 3.1 Constructing the Reference Distribution

We first examined the distribution of Pairwise Surface DSC scores in the validation cohort to identify an appropriate distribution family that it could be fit to. Candidate log-normal, log-*t*, and beta distributions were fitted to the validation scores, and Q-Q plots were generated using the fitted parameters (Figure S1 in the Supplementary Material). The log-*t* distribution produced the strongest linear agreement between observed and theoretical quantiles, and was therefore selected as the reference distribution. The final log-*t* fit had a location parameter of -5.48, a scale parameter of 0.692, and 3.45 degrees of freedom.

Pairwise DSC scores did not exhibit sufficient linear agreement with the theoretical quantiles of any of the candidate parametric distributions. Consequently, Pairwise DSC scores were transformed using ORQ normalization, which scaled the validation scores into a normal reference distribution for subsequent fitting and testing.

### 3.2 Risk Stratification Performance

To evaluate the proposed framework, significance scores were computed for each test segmentation using the reference distributions constructed from the validation cohort. Segmentations were then assigned to Low, Medium, or High Risk categories according to the predefined significance thresholds of 0.25 and 0.05.

The distributions of significance scores for the test cohort are shown in Figures S2 and S3 (Supplementary Material), while risk-level assignments are summarized in Table 1. Failed segmentations were assigned substantially lower significance scores than successful segmentations, supporting the use of significance scores as a proxy for segmentation quality. Consistent with this observation, the number of failure cases increased monotonically from the low risk to the high risk categories for both metrics, indicating that the proposed thresholds successfully stratified segmentations by risk (Table 1).

**Table 1.**
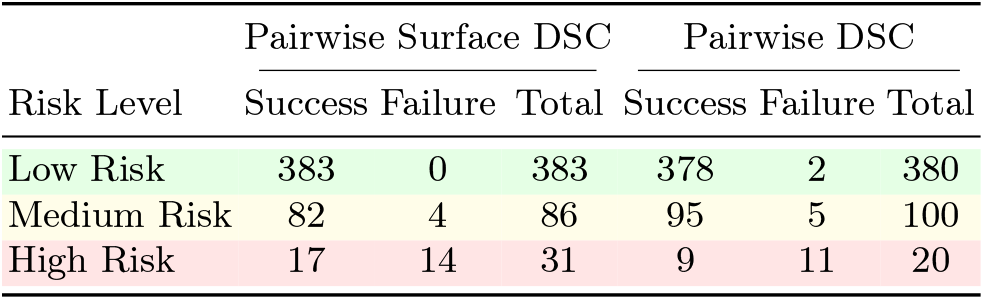
Risk-level assignments for successful and failed segmentations in the test cohort using Pairwise Surface DSC and Pairwise DSC.

Diagnostic performance at the Medium Risk and High Risk thresholds is summarized in Table 2. Pairwise Surface DSC generally outperformed Pairwise DSC, achieving perfect sensitivity at the Medium Risk threshold, while also achieving higher sensitivity at the High Risk threshold. Pairwise DSC achieved slightly higher specificity (0.98 versus 0.97) and positive predictive value (0.52 versus 0.45) at the High Risk threshold. Overall, the Medium Risk threshold prioritizes sensitivity for failure screening, whereas the High Risk threshold identifies a smaller subset of scans with a substantially higher probability of segmentation failure for high practitioner trust.

**Table 2.** Sensitivity, specificity, positive predictive value (PPV), and negative predictive value (NPV) for Pairwise Surface DSC and Pairwise DSC at each risk threshold. For the Medium Risk threshold, both Medium and High Risk segmentations were considered positive. Higher values indicate better performance. The metrics that each threshold was meant to target are shown in **bold**, and comparisons to the Youden’s index optimized cutoffs are shown.

|  | Risk Threshold | Sensitivity | Specificity | PPV | NPV |
| --- | --- | --- | --- | --- | --- |
| Pairwise Surface DSC | Medium Risk | **1.00** | 0.79 | 0.15 | **1.00** |
|  | High Risk | 0.78 | **0.96** | **0.45** | 0.99 |
|  | Youden’s Index | 0.89 | 0.89 | 0.23 | 1.00 |
| Pairwise DSC | Medium Risk | **0.89** | 0.78 | 0.13 | **0.99** |
|  | High Risk | 0.61 | **0.98** | **0.55** | 0.99 |
|  | Youden’s Index | 0.89 | 0.77 | 0.13 | 0.99 |

The Youden’s index optimized cutoffs fell between the Medium Risk and High Risk cutoffs in both cases, with the Surface DSC cutoff falling at a medium between the two and the DSC falling much closer to the Medium Risk cutoff. This resulted in metrics that were similarly spaced between those of the Medium Risk and High Risk cutoffs.

### 3.3 Impact of Failure Rate on Reference Distribution Modeling

The proposed framework assumes that the reference distribution is constructed primarily from successful segmentations, with failures comprising a relatively small fraction of the validation cohort. To evaluate the impact of violating this assumption, we simulated a range of failure rates by upsampling or downsampling the minority class and refitting the reference distribution for the Pairwise Surface DSC scores at each prevalence level. The resulting fitted distributions are shown in Figure 2 and a tabulation of the risk categories assigned to the test points by each reference distribution can also be found in Table 3.

**Fig. 2.**
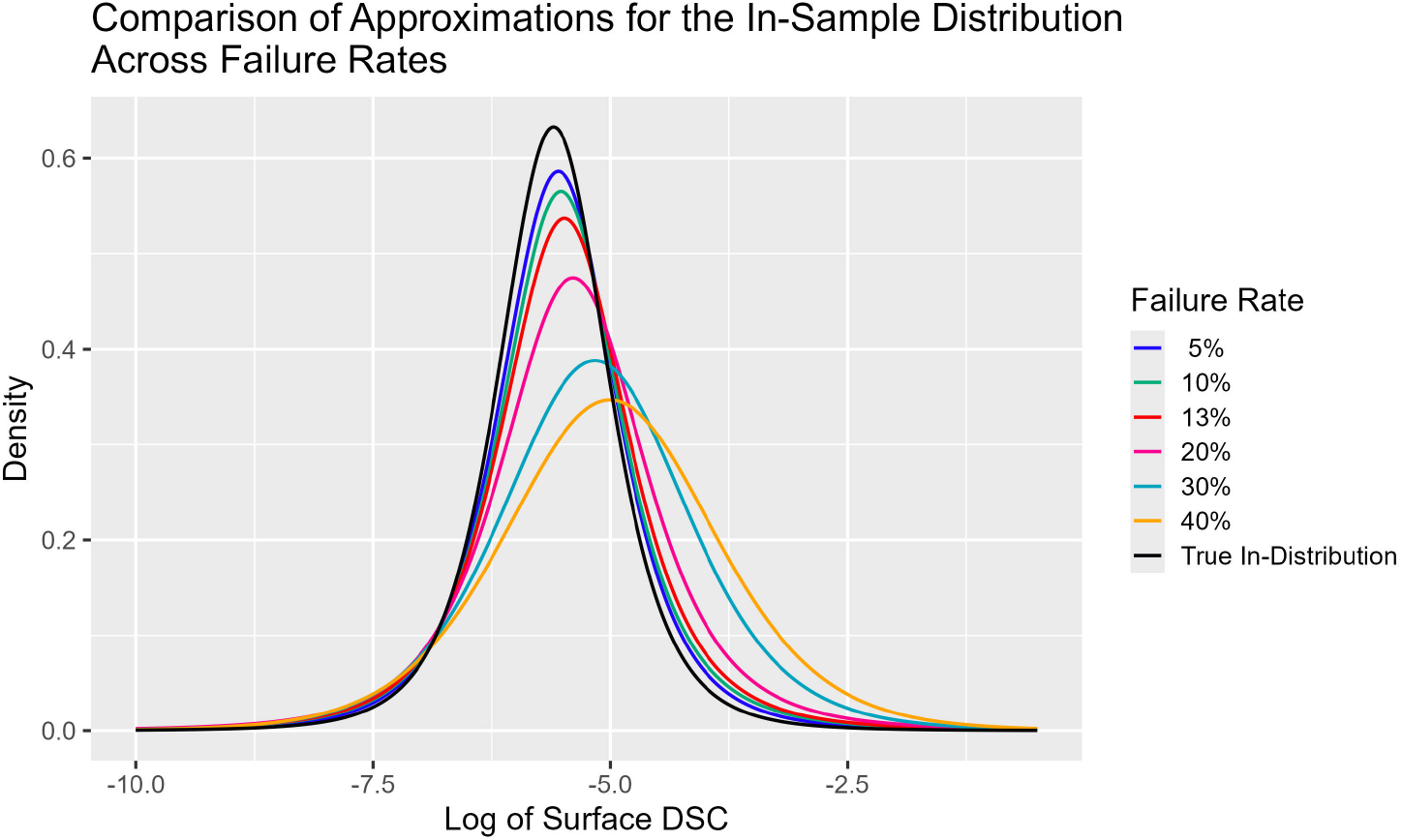
A comparison of the approximations of the in-distribution across different failure rates in the validation data.

**Table 3.** Counts of the successes and failures assigned to each of the three risk categories for a log-*t* in-distribution approximation approach on Surface DSC scores. Note that as the failure rate in the reference distribution increases, both successes and failures move towards the Low Risk category.

|  | Validation Set Failure Rate |  |  |  |  |  |  |  |  |  |
| --- | --- | --- | --- | --- | --- | --- | --- | --- | --- | --- |
|  | 5% |  | 10% |  | 20% |  | 30% |  | 40% |  |
|  | Success | Failure | Success | Failure | Success | Failure | Success | Failure | Success | Failure |
| Low Risk | 361 | 0 | 371 | 0 | 411 | 0 | 438 | 4 | 459 | 4 |
| Medium Risk | 98 | 4 | 92 | 4 | 64 | 7 | 40 | 5 | 22 | 7 |
| High Risk | 23 | 14 | 19 | 14 | 7 | 11 | 4 | 9 | 1 | 7 |

As the failure rate increases, the fitted reference distribution deviates progressively from the estimated true in-distribution. However, the approximation obtained at the observed validation failure rate of approximately 13% remains visually similar to the true in-distribution, and it can be seen in Figure 3 that it is at about 20% that our metrics start to suffer.

**Fig. 3.**
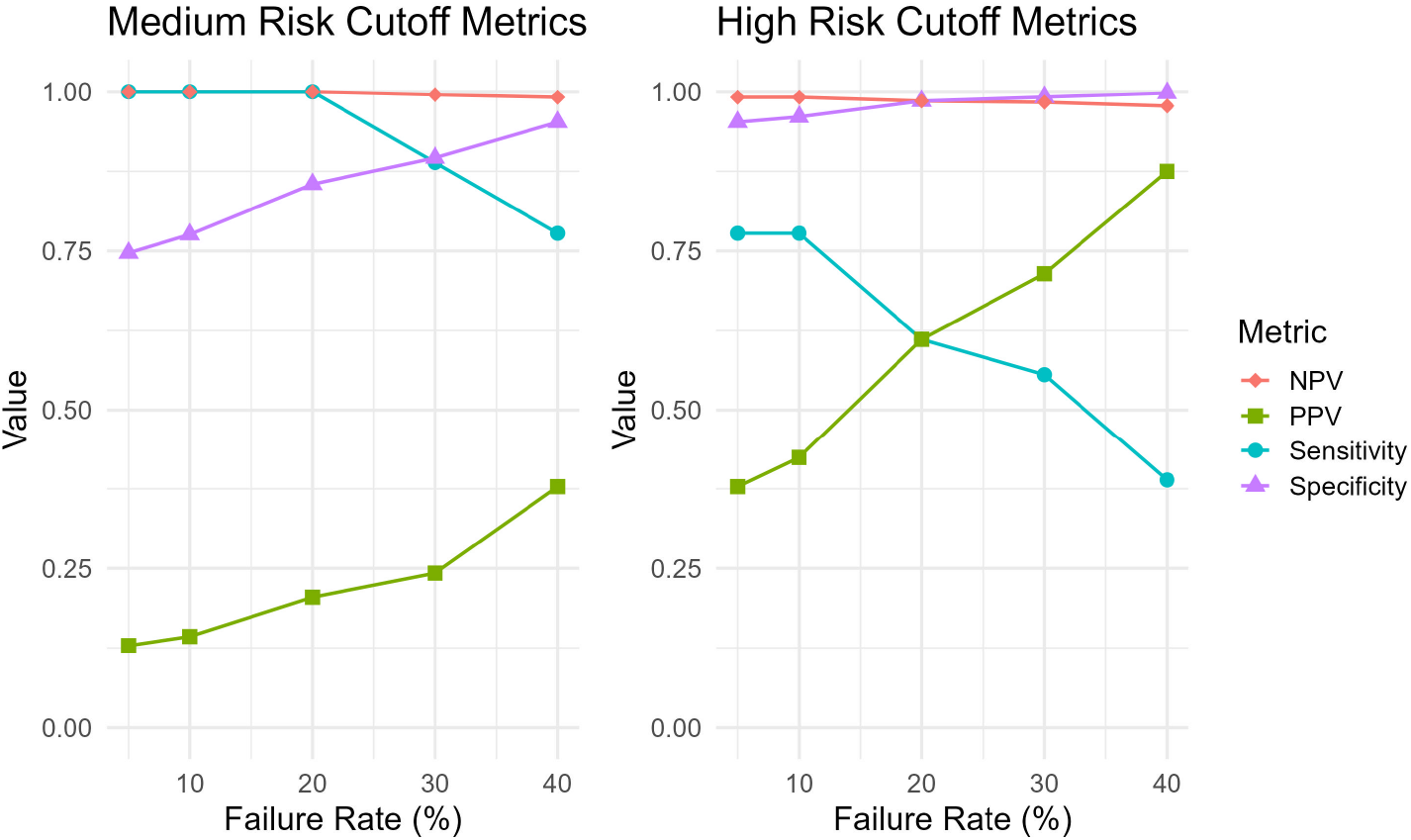
A visualization of how our performance metrics on the test set change given different failure rates in the validation set. With sensitivity as a priority, our process starts to lose its effectiveness at around a 20% failure rate. A failure rate of 40% pushes the Medium Risk cutoff to almost the exact same position as the High Risk cutoff at 5%.

### 3.4 Validation Sample Size

To determine the effectiveness of our validation set sample size, we used bootstrapping (R = 5000) to determine the variability of the location of our two cutoffs at that sample size. From our process, we estimated the Medium Risk cutoff for the Surface DSC score to be 0.9930163 and the High Risk cutoff to be 0.9805350. From the results of our bootsrapping, we are 95% confident that at a sample size of 400 the Surface DSC score for the Medium Risk cutoff will fall between 0.9920951 and 0.9937559 and the High Risk cutoff will fall between 0.9764717 and 0.9838607. These small margins are still very meaninful, but since even the lower bound for the Medium Risk cutoff still would’ve captured all of the failure cases in our test set we concluded that 400 was a sufficient validation set sample size.

## 4 Discussion and Conclusion

In this study, we introduced a framework for translating OOD scores into clinically interpretable risk levels without requiring labeled failures for calibration. Our Pairwise Surface DSC scores were well described by a log-*t* distribution and achieved strong risk stratification performance, with no failure cases assigned to the low risk category.

A key design goal of the proposed framework was to translate continuous OOD scores into actionable clinical guidance. We therefore adopted three risk categories rather than a binary classification scheme. The High Risk category is intended to identify a subset of segmentations with an elevated probability of failure, making review resources easier to prioritize, while the Medium Risk threshold is intended to provide high sensitivity so that problematic segmentations are unlikely to be overlooked. This dynamic makes our process preferable to using a single cutoff defined by optimizing the Youden’s J statistic which finds a happy medium between the two but does not quite share the strengths of either.

Regardless of the assigned risk level, automatic segmentations should still be reviewed before clinical use. The purpose of the proposed framework is not to replace human oversight but to provide additional information about the likelihood of segmentation failure. By making uncertainty explicit and clinically interpretable, the framework may help users calibrate their trust in automatic segmentations.

Because the proposed framework does not rely on labeled failures to assign risk levels at inference time, it remains dependent on several assumptions. First, the selected OOD score must exhibit a sufficiently consistent underlying distribution to permit construction of a reference distribution. In this study, Pairwise Surface DSC was well described by a log-*t* distribution, while Pairwise DSC required an ORQ transformation. Fortunately, this assumption can be assessed directly through Q-Q analysis, and alternative transformations may be applied when parametric distributions do not provide an adequate fit.

Second, the framework assumes a large reference cohort that is dominated by successful segmentations in order to accurately approximate the in-distribution. Our experiments suggest that the method is relatively robust to moderate contamination, even up to failure rates of 20 percent. The insight here is that while the approximation of the in-distribution from the validation dataset might change, that approximation doesn’t change the ordering of the points that are then assessed, where many of the failures lie far beyond the most extreme successes. This means that while some failures might get pushed from the High Risk to the Medium Risk or Low Risk categories, they will be followed by a large number of successes, keeping our confusion matrix statistics in a useful range.

Other work has found success in overcoming the challenge of a lack of failure cases using data augmentation, where models are trained on the actual data along with a population of synthetic points created to give examples for what OOD cases might look like [6,16]. While this might be effective in a variety of settings, it is unknown whether artificial failures would produce similar enough OOD scores to real failures to merit their use in threshold selection. In this study, we favored simply relying on the in-distribution to choose score thresholds, although future work towards creating artificial failure case points could hold promise.

Aside from providing clinically actionable guidance, the proposed framework may also be useful in retrospective studies where large numbers of segmentations are to be reviewed. Future work should investigate more robust methods for estimating the parameters of reference distributions in the presence of outliers and should extend our framework to additional anatomical structures, imaging modalities, and multiclass segmentation tasks. Overall, our findings demonstrate that OOD scores can be transformed into meaningful risk estimates without requiring labeled failures in the validation cohort and that Pairwise Surface DSC provides a promising foundation for risk stratification of liver CT segmentations.

## Data Availability

The MD Anderson data used in this study may be made available upon request in compliance with institutional IRB requirements and MD Anderson policies and guidelines.

## Acknowledgments

Research in this publication was supported by resources of the Image Guided Cancer Therapy Research Program at the University of Texas MD Anderson Cancer Center, the Tumor Measurement Initiative through the MD Anderson Strategic Initiative Development Program (STRIDE), and the National Cancer Institute of the National Institutes of Health under award numbers P30CA016672, 1R01CA221971, R01CA235564, and P01CA261669. N.S.S. was supported by the National Institutes of Health Image-Guided Cancer Therapies T32 Training Program Fellowship Grant (T32CA261856).

