## Supplemental Materials for "Label-Free Threshold Selection for Out-of-Distribution Detection in Liver CT Segmentation"

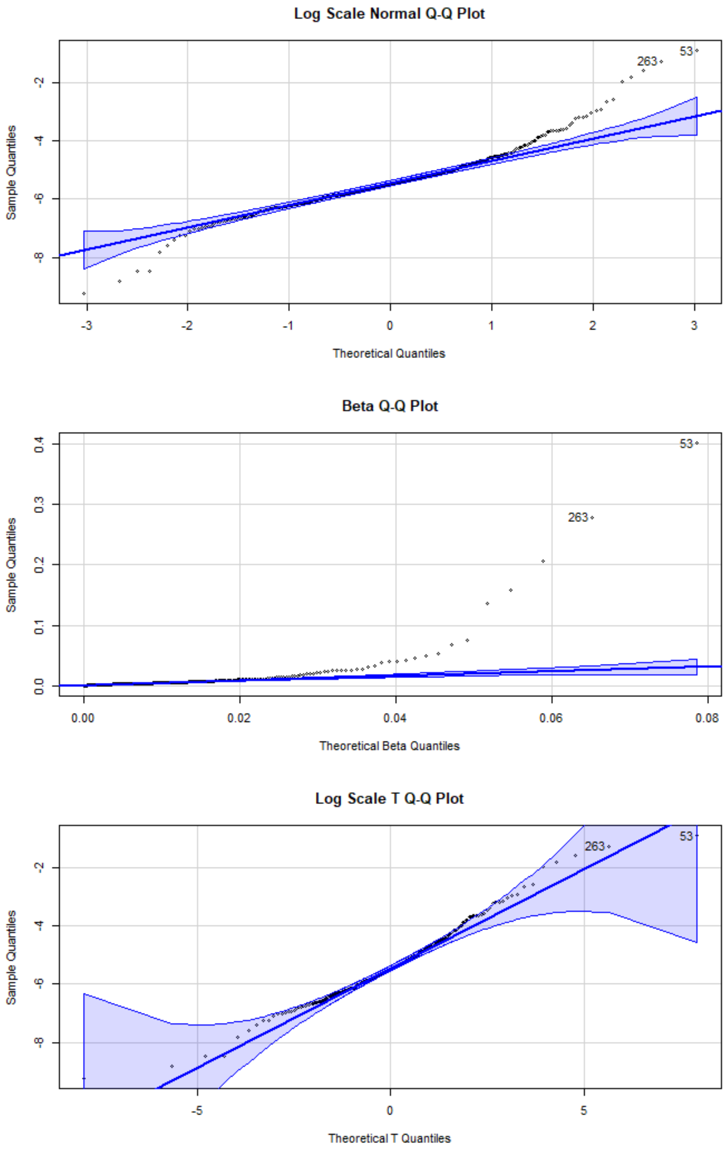


Figure S1: The Q-Q plots Pairwise Surface DSC scores fit to the log-normal, beta, and log-$t$ distribution families. In order to be considered a valid fit for each distribution, points must lie within the blue margins.

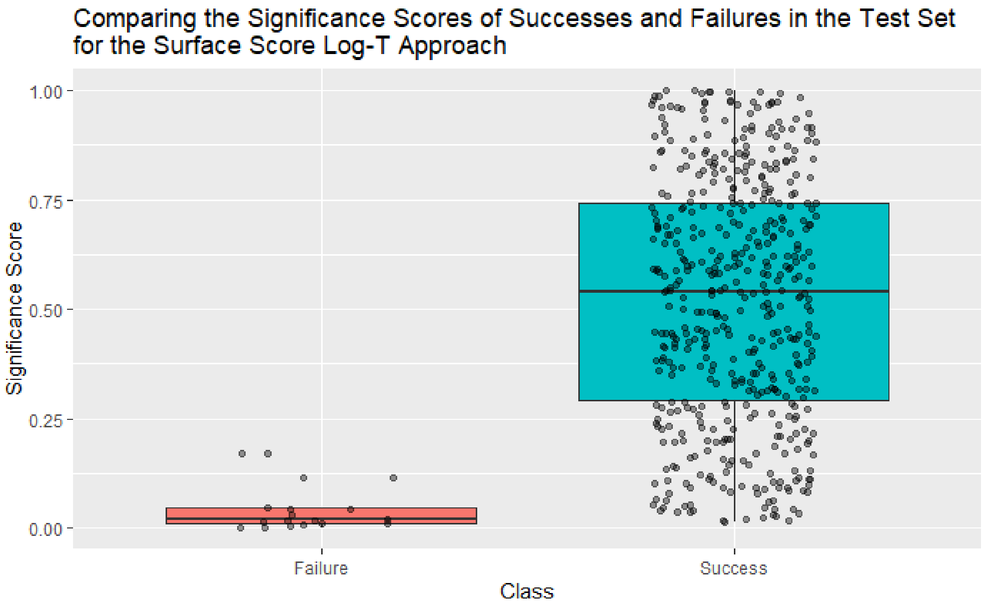


Figure S2: The significance scores assigned to each of the points in the test set for Surface DSC log-$t$ model. Points below 0.05 were flagged as “high risk”, between 0.05 and 0.25 were flagged as “medium risk”, and points above 0.25 were flagged as “low risk”.

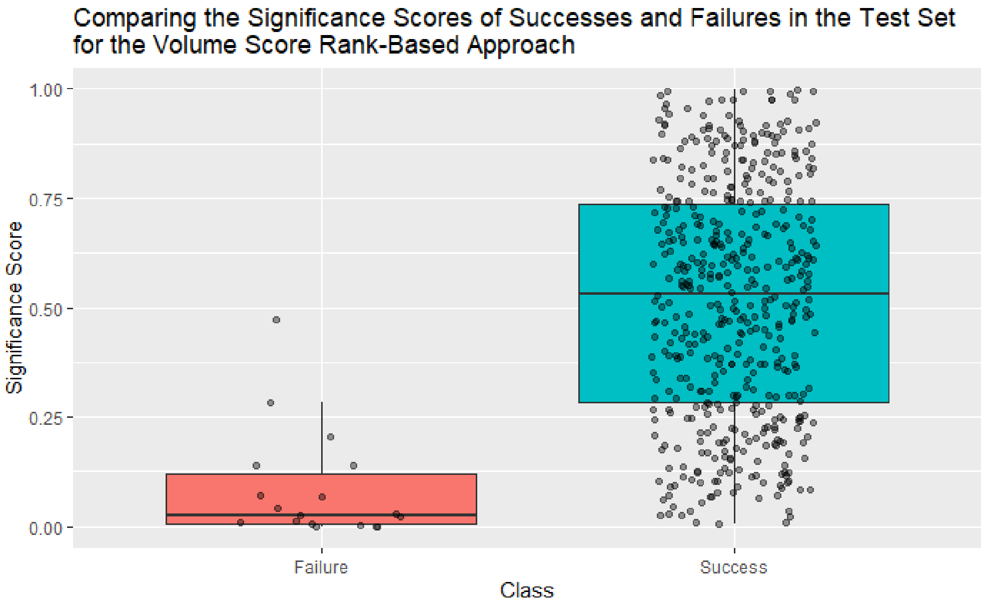


Figure S3: The significance scores assigned to each of the points in the test set for Volume DSC ORQ-normalized model. Points below 0.05 were flagged as “high risk”, between 0.05 and 0.25 were flagged as “medium risk”, and points above 0.25 were flagged as “low risk”.
